# Performance of urinary phenyl-γ-valerolactones as biomarkers of dietary flavan-3-ol exposure

**DOI:** 10.1101/2023.03.09.23287071

**Authors:** Benjamin H. Parmenter, Sujata Shinde, Kevin Croft, Kevin Murray, Catherine P. Bondonno, Angela Genoni, Claus T. Christophersen, Keren Biden, Colin Kay, Pedro Mena, Daniele Del Rio, Jonathan M. Hodgson, Nicola P. Bondonno

## Abstract

**Background:** Phenyl-*γ-*valerolactones (PVLs) have been proposed as potential biomarkers of dietary flavan-3-ol exposure.

**Objective:** We investigate the performance of a range of PVLs as biomarkers indicative of flavan-3-ol intake.

**Methods:** We report results of two companion studies: a 5-way randomised cross-over trial (RCT) and an observational cross-sectional study. In the RCT, 16 healthy participants were randomly assigned to 1-day flavan-3-ol rich interventions (of either apple, cocoa, black tea, green tea, or water [control]). Participants collected 24-hour urine and first morning urine samples, with diet standardised throughout. For each participant, one of the five intervention periods was randomly extended to two days, to monitor PVL kinetics following repeated days of flavan-3-ol exposure. In the cross-sectional study, 86 healthy participants collected 24-hour urines and concurrent weighed food diaries from which flavan-3-ol consumption was estimated using Phenol-Explorer. A panel of 10 urinary PVLs was quantified using liquid chromatography tandem mass spectrometry.

**Results:** In both studies, two urinary PVLs [5-(3’ s-hydroxyphenyl)-γ-valerolactone-4’-sulfate and tentatively identified 5-(4’-hydroxyphenyl)-γ-valerolactone-3’-glucuronide] were the principal compounds excreted (>75%). In the RCT, the sum of these PVLs was significantly higher than the water (control) following each intervention; individually, there was a shift from sulfation towards glucuronidation as the total excretion of PVLs increased across the different interventions. In the extended RCT intervention period, after two days of treatment, there was no evidence of accumulation of these compounds in the urine, and following withdrawal of treatment on the third day, there was a return towards negligible PVL excretion. All results were consistent, whether compounds were measured in 24-hour urine or first morning voids. In the observational study, the sum of the principal PVLs correlated dose-dependently (*R*_s_ = 0.37, *P* = 0.0006) with dietary flavan-3-ol intake, with similar associations for each individually.

**Conclusion:** Urinary 5-(3’-hydroxyphenyl)-γ-valerolactone-4’-sulfate and tentatively identified 5-(4’-hydroxyphenyl)-γ-valerolactone-3’-glucuronide are recommended biomarkers for dietary flavan-3-ol exposure.

## Introduction

Dietary flavan-3-ol intakes are associated with benefits for human health and disease (1). Found in foods and beverages such as tea, cocoa, red wine, nuts, grapes, apples and berries, flavan-3-ols are the subclass of flavonoids in the human diet with the highest intake (2). There are many different structures of flavan-3-ols, which range from single monomers (such as (epi)catechin, (epi)gallocatechin and their galloyl substituted derivatives) to polymers, or chains of two or more molecules (referred to as proanthocyanidins) (2). Following their consumption, most flavan-3-ols reach the intestine, where they undergo microbial metabolism leading to the formation of specific metabolites, including phenyl-*γ-*valerolactones (PVLs) (2). Once absorbed, PVLs may be further metabolised to glucuronidated, methylated or sulfated forms, giving rise to a wide range of PVL structures (2). As PVLs form exclusively from flavan-3-ol metabolism, show a dose-response relationship with intake, and a half-life potentially sufficient to produce steady-state like conditions with regular flavan-3-ol consumption, PVLs have been proposed as candidate biomarkers for dietary flavan-3-ol exposure (3).

Certain PVLs may be suitable as objective biomarkers of flavan-3-ol exposure, with respect to intake, following metabolism. Validated PVLs biomarkers could then be applied to evaluate diet-disease relationships in epidemiological studies, circumventing many limitations associated with subjective dietary assessment, thereby potentially improving upon risk estimates obtain from association studies (4). However, a limited number of studies have evaluated the performance of PVLs as possible biomarkers for dietary flavan-3-ol exposure, hindering their application to epidemiologic investigations. Therefore, in order to evaluate the suitability of urinary PVLs as biomarkers, in this investigation, we aimed to: 1) identify PVL biomarkers that reflect intakes of commonly consumed flavan-3-ol rich foods (i.e., apples, cocoa, black tea and green tea), 2) as 24-hour urine samples are rarely collected in epidemiological studies, we sought to determine whether PVLs measured in 24-hour urines and first morning voids analogously reflect flavan-3-ol exposure, 3) evaluate PVL kinetics (in both 24-hour urine and first morning void samples), following repeated days of flavan-3-ol intake, to examine how changes in habitual dietary intake may be reflected in the urine and 4) apply the biomarkers to a free-living population, as a proof of concept, to determine whether urinary PVLs rank flavan-3-ol exposure in a dose-dependent manner among those consuming their regular diet. In this way, we undertook a thorough evaluation of the performance of PVLs as potential biomarkers for dietary flavan-3-ol exposure.

## Methods

We report two companion studies: a clinical trial and an observational investigation. The clinical trial was approved by the University of Western Australia Human Research Ethics Committee (RA/4/20/5366). The RCT was prospectively registered with the Australian New Zealand Clinical trials registry (ACTRN12619001553167) under the World Health Organisation, Universal Trial Number: U1111-1236-7988. The observational study was approved by the Edith Cowan University, Human Research Ethics Committee (13402). The conduct and reporting of the clinical trial adheres to the Consolidated Reporting Statement for Randomised Controlled Trials (CONSORT). The observational study adheres to the reporting recommendations of the STrengthening the Reporting of OBservational studies in Epidemiology (STROBE) statement. All participants provided written informed consent before inclusion in each study.

### Clinical trial

A 5-way randomised controlled cross-over trial was conducted to: 1) identify PVL biomarkers that reflect intakes of commonly consumed flavan-3-ol rich foods (i.e., apple, cocoa, black tea, and green tea), and 2) as 24-hour urine samples are rarely collected in epidemiological studies, we sought to determine whether PVLs measured in 24-hour urine and first morning void samples analogously reflect flavan-3-ol exposure. Healthy participants (*n* = 16 completed the trail) were recruited from the Perth and Peel regional area, Australia, the details of which can be found in the Online Supplement. Participants received, in a random order, five 1-day (3 times/day) flavan-3-ol test interventions: 1) 250 ml black tea (Nerada brand), 1 tea bag steeped for two minutes in freshly boiled water (no milk, sugar permitted, served with the tea bag removed after squeezing till drop-less to extrude flavan-3-ols); 2) 250 ml green tea (Nerada brand), 1 tea bag steeped for two minutes in freshly boiled water (no milk, sugar permitted, served with the tea bag removed after squeezing till drop-less to extrude flavan-3-ols); 3) a Cripps Pink apple marketed as Pink Lady TM (skin on [mean ± SD weight of intervention apples: 539.4 ± 25.6 g/d/person]); 4) a cocoa beverage (CocoaVia™ Daily Cocoa Extract Supplement, 450 mg cocoa flavanol powder sachet) served in hot water (no milk, sugar permitted) and; 5) 250 ml of water (control). To standardise intervention intake, participants consumed the test products at 9:00 am, 12:00 pm and 3:00 pm. To assist with compliance, participants received text messages at these times, to remind them to consume the intervention. To monitor adherence, participants kept food diaries on the days of the intervention, as well as the preceding day to record timings of the interventional intake as well as all other food and fluid. A qualified dietitian met with the participants following each intervention period and cross-checked the food diary against participant responses. Each intervention period was preceded by a 1-day low flavan-3-ol diet, and each subsequent intervention period was separated by a >4-day washout period. All volunteers were provided with all meals for the intervention days as well as the meal the night before. These meals were prepared using foods that contain no or very low flavan-3-ol contents by the Nutrition Laboratory at Edith Cowan University; full details can be found in the Online Supplement. Within the RCT, for each participant, one of the five intervention periods was randomly extended to two days, to monitor PVL kinetics following repeated days of flavan-3-ol exposure, to examine how changes in habitual dietary intake, may be reflected in the urine (**Supplementary Figure 1**). For logistical reasons this long intervention period was conducted at the first visit, with all remaining periods being single day interventions. In an attempt to allocate a balanced number of individuals to receive each treatment in each intervention period, each individual was randomly allocated a treatment sequence using Williams type latin squares. From this design a series of twenty sequence orders were produced, in random succession, by a researcher not involved in the study, which were assigned to individuals in the order they were enrolled. The investigator (BHP) who enrolled participants had no knowledge of the sequence orders, ensuring concealment allocation. Given the nature of the interventions, blinding was not possible although, given our objective outcomes, this is unlikely to affect our results. On the days of the interventions, participants collected 24-hour urines which were stored in portable cooler boxes containing ice packs. To standardise timing of the 24-hour urine collection, against timing of the interventions across periods, participants voided their bladder at 8.55 am, and thereafter began their 24-hour urine collections in each period. During the extended intervention period, participants collected consecutive 24-hour urine samples for 3 days in addition to first morning voids, the timings of which are detailed in Supplementary Figure 1.

### Observational study

An observational investigation was conducted to examine whether the excretion of urinary PVLs reflect flavan-3-ol exposure in a dose-dependent manner among a free-living population. For this investigation, we used data and stored samples (collected within <6 years, stored at −80 °C and not subject to prior freeze thaw events) from a previously published study (5). The original investigation compared gastrointestinal health across subjects with long-term (> 1 year) adherence to a Paleolithic diet to those following a standard Australian diet (5). Briefly, healthy participants (*n* = 86 with complete data) were recruited to a cross-sectional study, collecting 24-hour urine samples and concurrent weighed food diaries. Intake of macronutrients were estimated using the AUSNUT 2007 database. Consumption of flavonoids were estimated using the (poly)phenol food-composition database published by Knaze *et al*., (6) which derives original content values from Phenol-Explorer (7). This database extends on Phenol-Explorer and provides information on 437 polyphenols in 19,899 generic and multi-ingredient food items computed using yield and retention factors (where applicable) to account for food preparation methods (6). The database provides flavonoid intake values without previous hydrolysis of glycosides or esters, except for proanthocyanidins (trimers to polymers), for which normal phase high performance liquid chromatography is used. The food diaries showed participants reported consumption of >350 unique foods and beverages in addition to >120 recipes, consumed across >600 eating occasions. Foods and beverages in the diaries were manually matched based on the food name, processing, and preparation method, to the most similar food reported in the food-composition database by a qualified dietitian. Multi-ingredient recipes not found in the food-composition database were broken down into single ingredients before matching to specific foods. Daily flavan-3-ol intake (mg/day) was calculated for each study participant by multiplying the flavonoid content reported for each food, by the weight reported in the food diary.

### Analysis of urinary phenyl-γ-valerolactones

A panel of 10 PVLs were quantified in all urine samples using liquid chromatography tandem mass spectrometry (LC-MS/MS). Of the 10 measured PVLs, three were mono-hydroxylated PVLs including: 5-phenyl-γ-valerolactone-3’
s-sulfate (PVL-3’-Sulf), 5-phenyl-γ-valerolactone-4’-sulfate (PVL-4’-Sulf) and 5-phenyl-γ-valerolactone-3’-glucuronide (PVL-3’-Gluc); while seven were di-hydroxylated PVLs including: 5-(3’,4’-Dihydroxyphenyl)-γ-valerolactone (3’,4’-DiOH-PVL), 5-(3’-hydroxyphenyl)-γ-valerolactone-4’-sulfate (3’-OH-PVL-4’-Sulf), 5-(4’-hydroxyphenyl)-γ-valerolactone-3’-glucuronide (4’-OH-PVL-3’-Gluc), 5-phenyl-γ-valerolactone-3’,4’-disulfate (PVL-3’,4’-DiSulf), 5-phenyl-γ-valerolactone-3’-sulfate-4’-glucuronide (PVL-3’-Sulf-4’-Gluc), 5-(4’-hydroxyphenyl)-γ-valerolactone-3’-methoxy (4’-OH-PVL-3’-Methoxy), and 5-(5’-hydroxyphenyl)-γ-valerolactone-3’-glucuronide (5’-OH-PVL-3’-Gluc). Of these compounds, five were tentatively identified, without the use of reference standards (including: PVL-3’-Sulf, 4’-OH-PVL-3’-Gluc, PVL-3’,4’-DiSulf, PVL-3’-Sulf-4’-Gluc and 4’-OH-PVL-3’-Methoxy). Details of the full methodology in addition to structural information and nomenclature used to denote the PVLs reported throughout this manuscript is detailed in the Online Supplement (**Supplementary Tables 1 and 2, and Supplementary Figure 2**).

### Statistical analyses

All analyses were performed using R Statistical Software (8). For analysis of the RCT, differences in total (i.e., summed quantities of) PVL excretion across interventions was assessed by linear mixed model, with total PVL concentration as the outcome, intervention as a fixed effect, and subject as an intercept only random effect. Differences among all individual PVLs were assessed in a single linear mixed model, with PVL concentration as the outcome, intervention and PVL type as fixed effects with an interaction, and subject as an intercept only random effect. In the extended RCT intervention period, where changes in PVL excretion following repeated days of flavan-3-ol containing interventions was of interest, differences in total (i.e., summed quantities of) PVL excretion was assessed by linear mixed model, with total PVL concentration as the outcome, intervention and day as fixed effects with an interaction, and subject as an intercept only random effect. Differences among all individual PVLs were assessed in a single linear mixed model, with PVL concentration as the outcome, intervention, PVL type and day as fixed effects with a 3-way interaction, and subject as an intercept only random effect. For normalization purposes, for analysis of the RCT, all outcome variables were assessed as log(x+1) throughout statistical analysis, although all figures and descriptive statistics are presented using data without transformation. Correlations between variables were performed using Spearman’s correlation coefficient (*R*_s_) and repeated measures correlations (*R*_m_) where appropriate (9). Two-tailed *P* < 0.01 was considered significant.

## Results

### Clinical trial

Recruitment began on 16 February 2021 and the study ended on 10 June 2021. Of the 75 participants screened, 18 were randomly assigned, 16 of whom completed the clinical trial (**Figure 1**). The two participants who withdrew after randomization did so due to medical reasons unrelated to the trial. Participants had a median ± SD age of 67.0 ± 6.1 years, were within a healthy weight range [20.2–30.6 kg/m^2^], and roughly half were females (56.2%; **Table 1**). Most participants were Caucasian (81.3%), none took prescription medication, and the majority had never smoked (81.3%). Analysis of the 24-hour urine showed that two PVLs [3’ s-OH-PVL-4’-Sulf and tentatively identified 4’-OH-PVL-3’-Gluc] were the principal compounds excreted, constituting ≥75% of the measured metabolites following each intervention except green tea, wherein they comprised ∼62% of excreted metabolites. The sum of these PVLs was significantly higher than the water (control) following each intervention; individually, there was a shift from sulfation towards glucuronidation as the total excretion of PVLs increased across the different test products (black tea < apple < green tea < cocoa) (**Figure 2, Supplementary Table 3**). Several other PVLs were also excreted in significantly higher concentrations following specific interventions, although, each of these compounds were excreted in minor to moderate concentrations (Figure 2). In the extended RCT intervention period, when we examined PVL excretion following repeated days of flavan-3-ol intake, we found no evidence of accumulation of 24-hour urinary PVLs (after two days of treatment) and following withdrawal of treatment (on the third day), there was a significant return towards negligible PVL excretion (**Figure 3**; **Supplementary Table 4**). Analysis of the first morning spot urine samples showed the pattern of PVL excretion almost mirrored findings from the 24-hour urine analysis (**Figure 4**; **Figure 5**; **Supplementary Table 5; Supplementary Table 6**); the correlation between first morning voids and 24-hour urine PVL status was ≥0.85 for the compounds recovered in the highest abundance (**Figure 6**).

**Figure 1.**
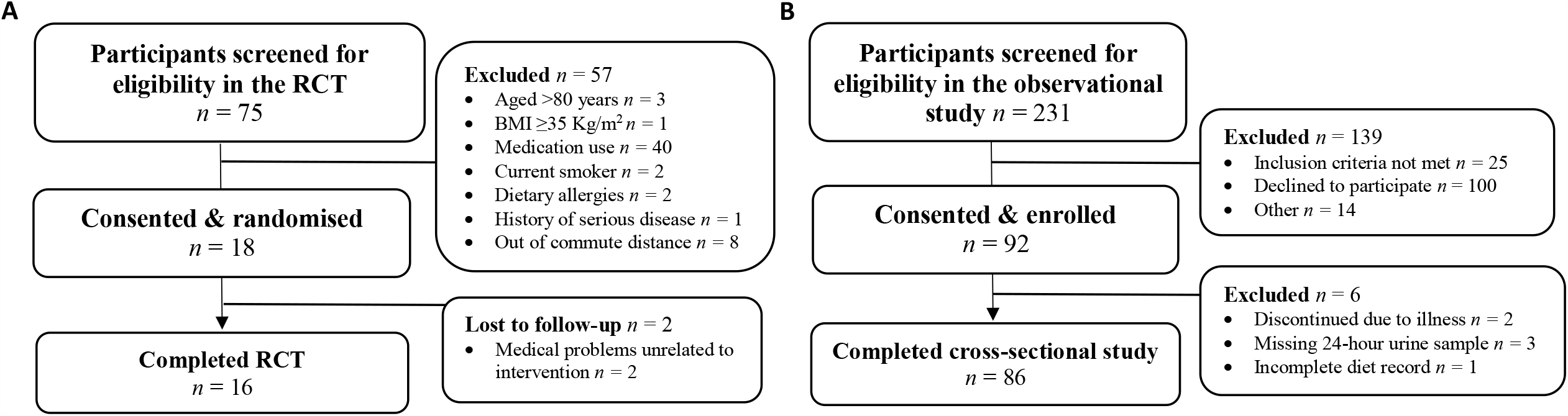
Participant recruitment and flow through the clinical trial (Panel A) and the observational study (Panel B).

**Table 1.** Characteristics of participants in the clinical trial^*1*^

|  | All participants<br>(n = 16) |
| --- | --- |
| **Demographic** |  |
| Sex [male; n (%)] | 7 (43.8) |
| Age (y) | 67.0 ± 6.1 |
| Education [n (%)] |  |
| High school or less | 4 (25) |
| Diploma or trade | 2 (12.5) |
| Bachelor degree or above | 10 (62.5) |
| Occupation [n (%)] |  |
| Full-time | 1 (6.25) |
| Part-time | 2 (12.5) |
| Retired | 13 (81.3) |
| Ethnic background [n (%)] |  |
| Caucasian | 13 (81.3) |
| Asian | 3 (18.7) |
| Smoking status [n (%)] |  |
| Current | 0 (0) |
| Former | 3 (18.7) |
| Never | 13 (81.3) |
| Vigorous exercise (MET-mins/week) | 2916.3 ± 1547.7 |
| Basal metabolic rate (MJ/d) | 5934 ± 1287.4 |
| **Anthropometry** |  |
| Body mass index (kg/m<sup>2</sup>) | 24.3 [22.1–27.0] |
| Hip-to-waist ratio |  |
| Males (healthy WHR: ≤0.95) | 0.91 ± 0.06 |
| Females (healthy WHR: ≤0.80) | 0.81 ± 0.09 |
| Body fat (%) |  |
| Males | 23.7 ± 3.1 |
| Females | 30.4 ± 9.1 |
| **Clinical characteristic** |  |
| Systolic blood pressure (mmHg) | 130.1 ± 15.6 |
| Diastolic blood pressure (mmHg) | 72.4 ± 9.6 |
| Mean arterial pressure (mmHg) | 95.1 ± 11.4 |
| Heart rate (beats/min) | 63.1 ± 7.8 |

<sup>I</sup>Values are means ± SDs or medians [IQRs] unless otherwise indicated. WHR, waist to hip ratio.

**Figure 2.**
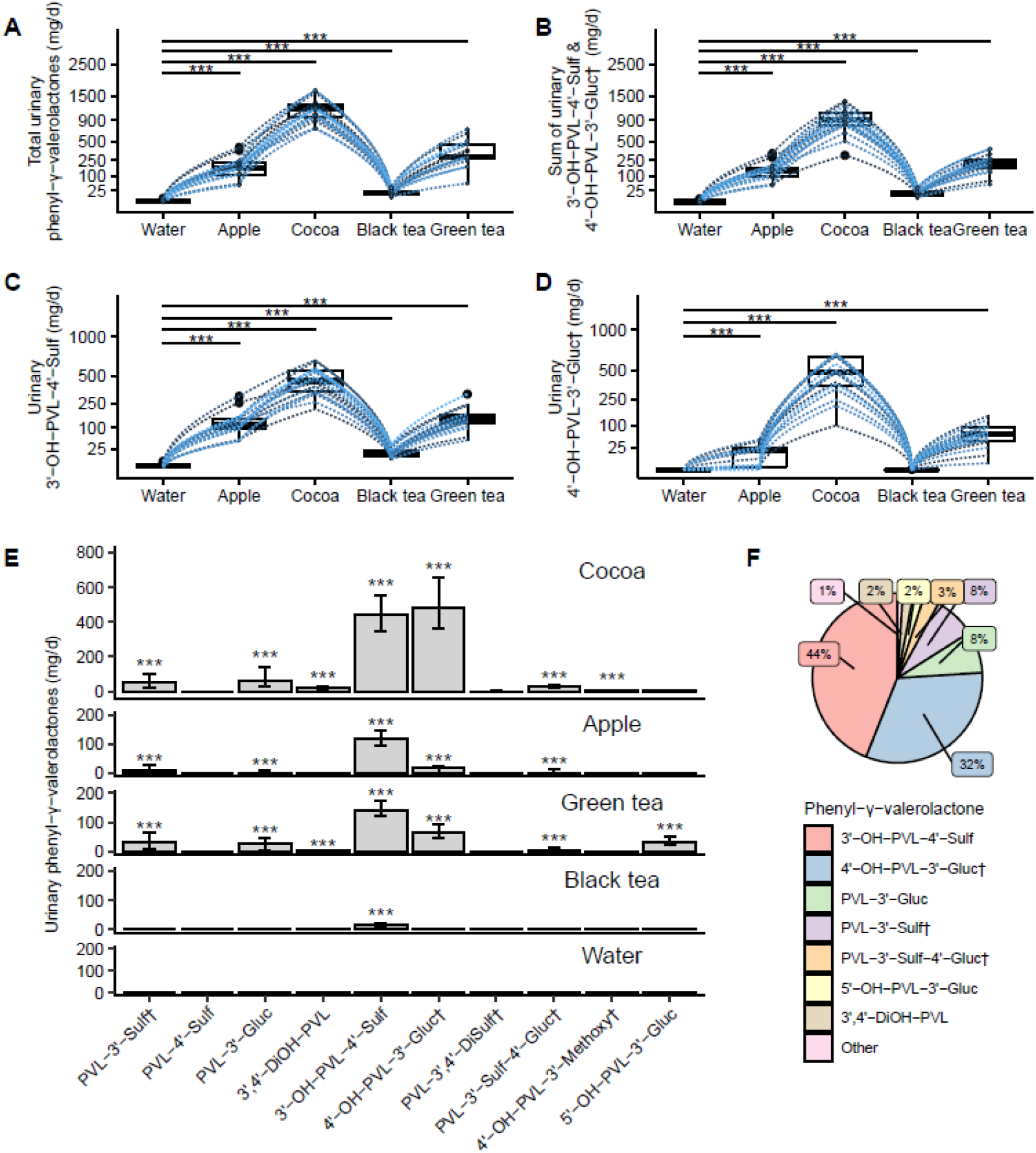
Excretion of phenyl-γ-valerolactones in 24-hour urine following flavan-3-ol containing interventions of either cocoa, apple, green tea, black tea or a water control in a 5-way randomised cross over trial (*n* = 16). Panels **A** through **D** show box and whisker plots of individual participant data for selected phenyl-γ-valerolactones of interest. Panel **E** displays the median [IQR] excretion of each measured phenyl-γ-valerolactone, with stars highlighting significantly higher excretion of specific phenyl-γ-valerolactones, after intake of the different interventions, in comparison to the water control. Panel **F** shows the average percent excretion of the measured phenyl-γ-valerolactones across interventions. Differences in total phenyl-γ-valerolactone excretion across interventions was assessed by linear mixed model on log-transformed data, when applicable. \**P* ≤ 0.01, \*\**P* ≤ 0.001, \*\*\**P* ≤ 0.0001. Figures presented without data transformation. Descriptive statistics of this analysis can be found in Supplementary Table 3. Structural information about the phenyl-γ-valerolactones is detailed in Supplementary Table 1. ^†^Tentatively identified compound.

**Figure 3.**
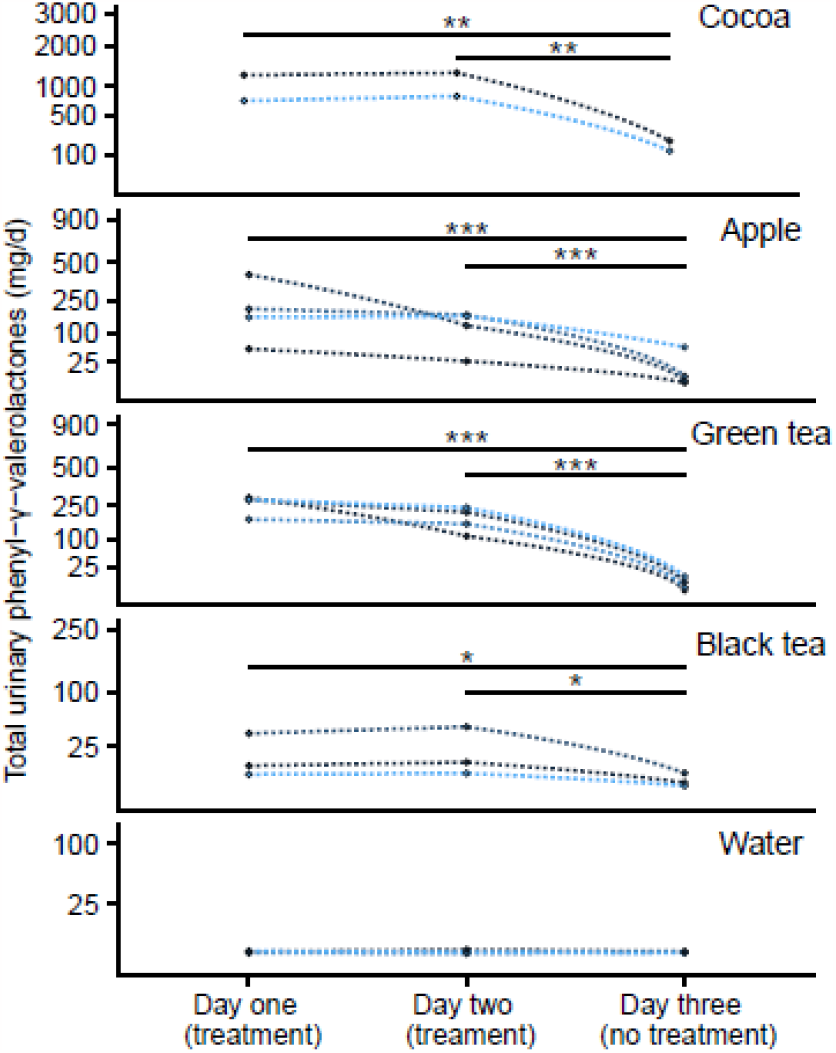
Excretion of total phenyl-γ-valerolactones in 24-hour urine following repeated days of flavan-3-ol containing interventions (on days one and two), preceding withdrawal of treatment (on the third day), collected during the extended RCT intervention period (cocoa *n* = 2; apple *n* = 4; green tea *n* = 3; black tea *n* = 3; water *n* = 3). Differences in total phenyl-γ-valerolactone excretion across days was assessed by linear mixed model on log-transformed data, when applicable. Stars highlight significantly different excretion of total phenyl-γ-valerolactones between days. \**P* ≤ 0.01, \*\**P* ≤ 0.001, \*\*\**P* ≤ 0.0001. Figures presented without data transformation. Descriptive statistics of this analysis can be found in Supplementary Table 4.

**Figure 4.**
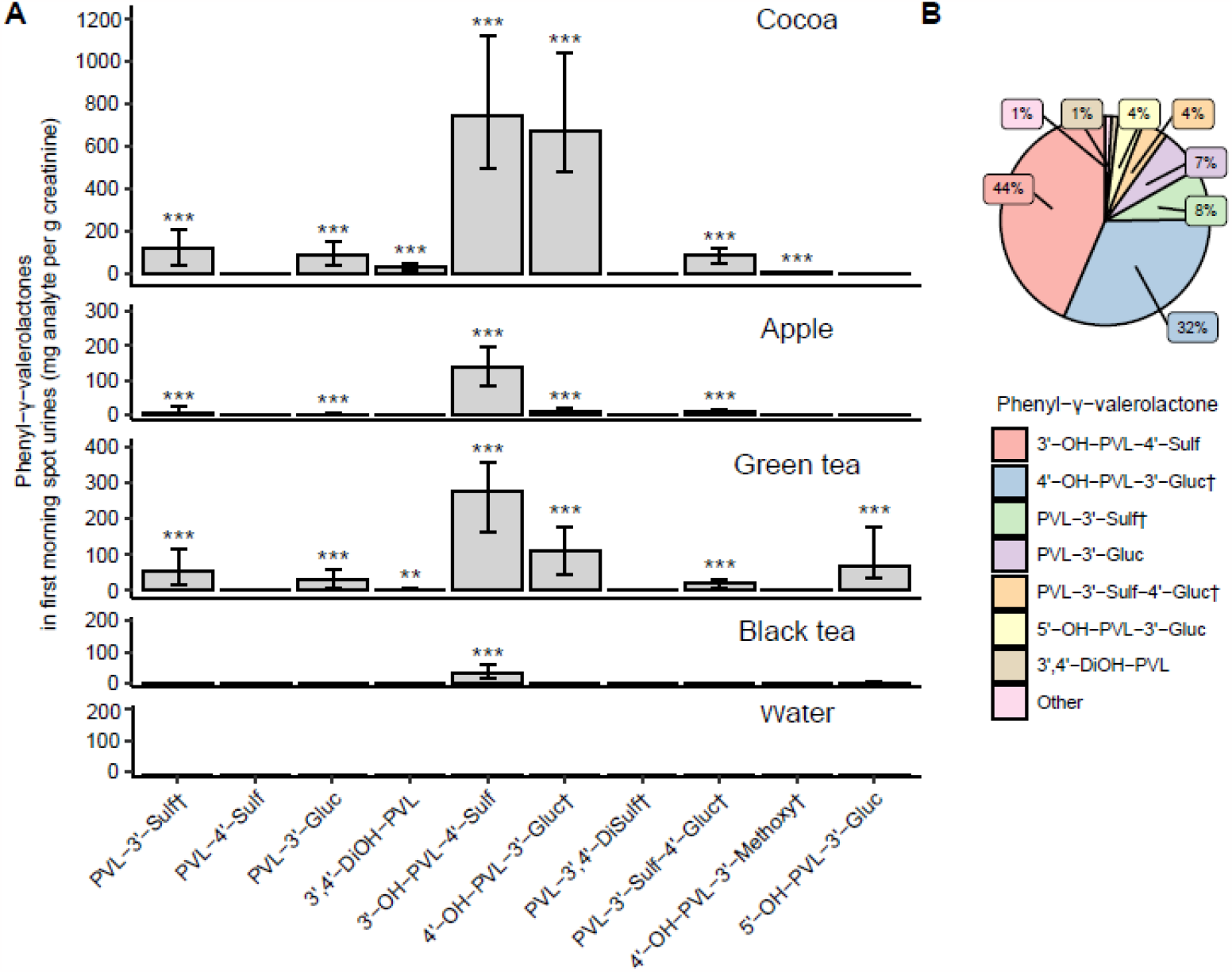
Excretion of phenyl-γ-valerolactones in first morning spot urine voids following flavan-3-ol containing interventions (consumed on the previous day) of either cocoa, apple, green tea, black tea or a water control in a 5-way randomised cross over trial (*n* = 16). Panel **A** displays the median [IQR] excretion of each measured phenyl-γ-valerolactone, with stars highlighting significantly higher excretion of specific phenyl-γ-valerolactones, after intake of the different interventions, in comparison to the water control. Panel **B** shows the average percent excretion of the measured phenyl-γ-valerolactones across interventions in the spot samples. Differences in total phenyl-γ-valerolactone excretion across interventions was assessed by linear mixed model on log-transformed data, when applicable. \**P* ≤ 0.01, \*\**P* ≤ 0.001, \*\*\**P* ≤ 0.0001. Figures presented without data transformation. Descriptive statistics of this analysis can be found in Supplementary Table 5. Structural information about the phenyl-γ-valerolactones is detailed in Supplementary Table 1. ^†^Tentatively identified compound.

**Figure 5.**
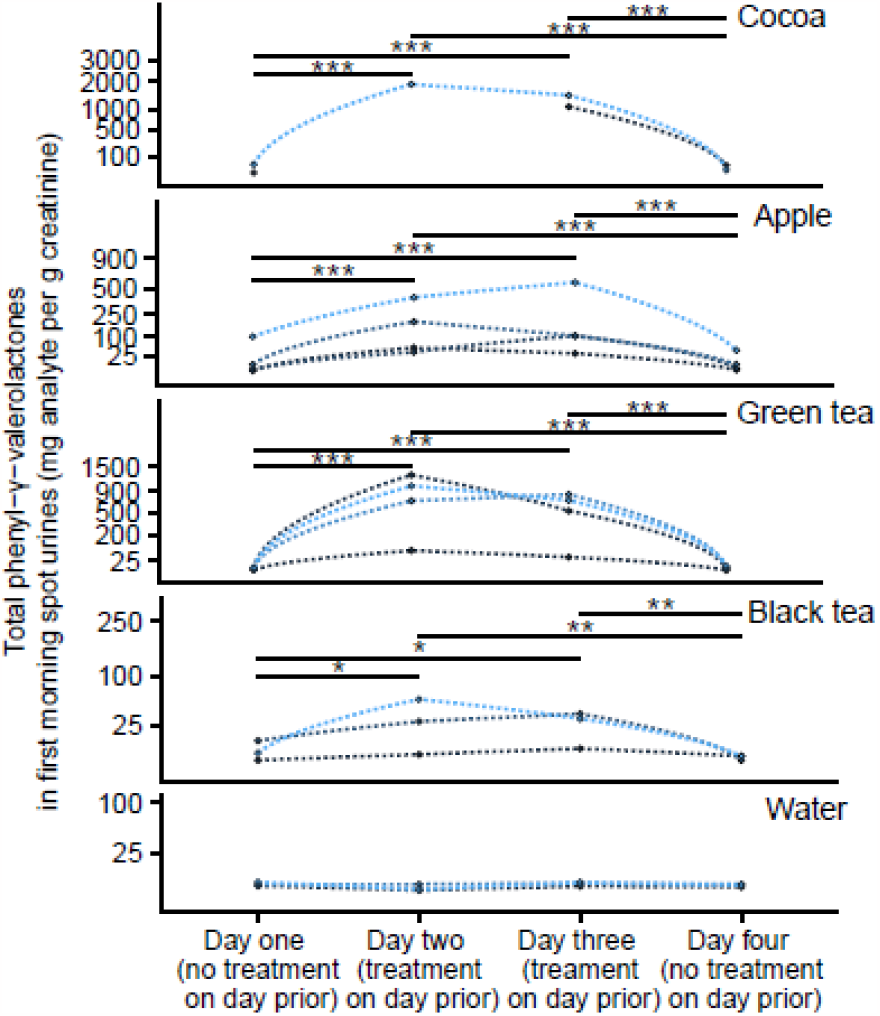
Excretion of phenyl-γ-valerolactones in first morning spot urine samples following repeated days of flavan-3-ol containing interventions collected during the extended RCT intervention period (cocoa *n* = 2; apple *n* = 4; green tea *n* = 3; black tea *n* = 3; water *n* = 3). Differences in total phenyl-γ-valerolactone excretion across days was assessed by linear mixed model on log-transformed data, when applicable. Stars highlight significantly different excretion of total phenyl-γ-valerolactones between days. \**P* ≤ 0.01, \*\**P* ≤ 0.001, \*\*\**P* ≤ 0.0001. Figures presented without data transformation. Descriptive statistics of this analysis can be found in Supplementary Table 6. Participants with missing data are shown as disconnected lines.

**Figure 6.**
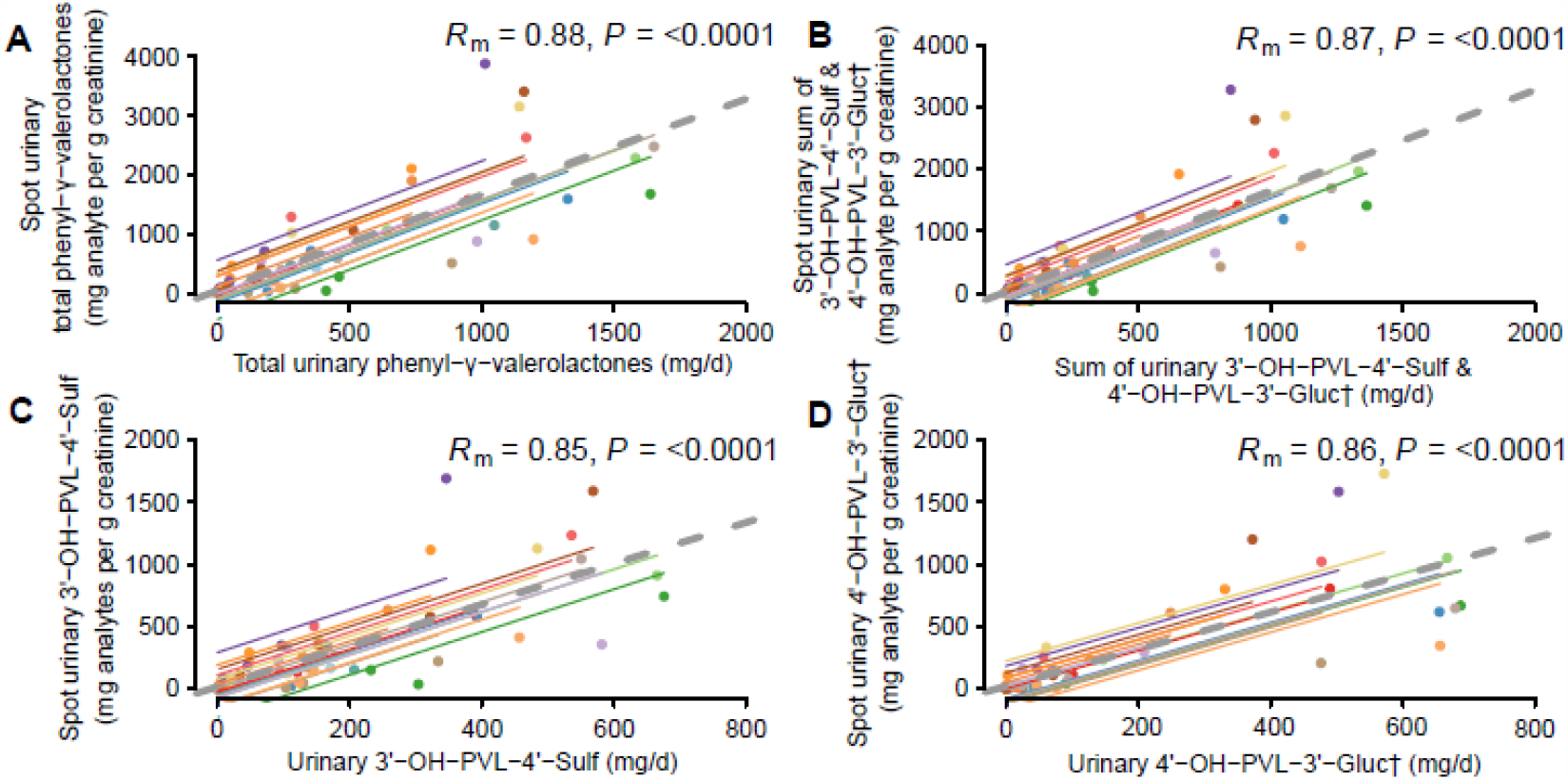
Relation between phenyl-γ-valerolactones excreted in 24-hour urines and first morning voids collected throughout a 5-way randomised cross over trial (*n* = 16). All correlations were calculated using repeated measures correlation (*R*_*m*_). In the figures, observations from the same participant are given the same color, with corresponding lines to show the repeated measures fit for each participant. Structural information about the phenyl-γ-valerolactones is detailed in Supplementary Table 1. ^†^Tentatively identified compound.

### Observational study

Of the 231 participants screened, 92 enrolled to the observational study, of which, complete data was available on 86 individuals (Figure 1). Participants were aged 20 to 69 years, most were females (75.6%), had a BMI ≤30 km/m^2^ (90.1%), and none were current smokers (**Table 2**). The range of flavan-3-ol intake (excluding theaflavins and thearubigins) was wide (median [IQR], 290 [78.3–460.3] mg/day) as was 24-hour urinary excretion of total PVLs (median [IQR], 135 [53.5–283.2] mg/day; **Supplementary Table 7**). Both 3’
s-OH-PVL-4’-Sulf and tentatively identified 4’-OH-PVL-3’-Gluc were the top two metabolites excreted in the observational study, constituting ≥75% of the PVLs recovered in the urine, although, 3’-OH-PVL-4’-Sulf accounted for a greater proportion (68%) in comparison to 4’-OH-PVL-3’-Gluc (10%) (**Figure 7**). The sum of these correlated dose-dependently with total dietary flavan-3-ol intake (*R*_s_ = 0.37, *P* = 0.0006), with similar associations for each compound individually (3’-OH-PVL-4’-Sulf, *R*_s_ = 0.35, *P* = 0.001; 4’-OH-PVL-3’-Gluc, *R*_s_ = 0.33, *P* = 0.002) (Figure 7).

**Table 2.**
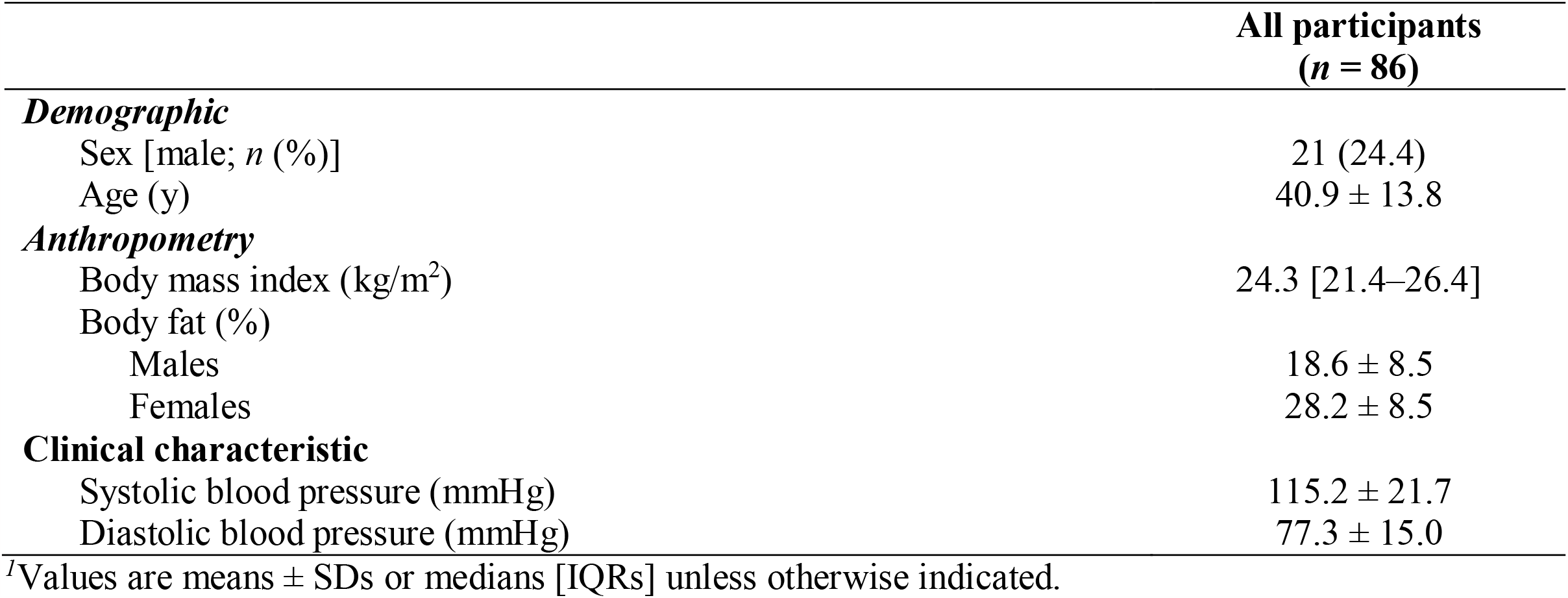
Characteristics of participants in the observational study^*1*^

|  | All participants<br>(*n* = 86) |
| --- | --- |
| ***Demographic*** |  |
| Sex [male; *n* (%)] | 21 (24.4) |
| Age (y) | 40.9 ± 13.8 |
| ***Anthropometry*** |  |
| Body mass index (kg/m<sup>2</sup>) | 24.3 [21.4–26.4] |
| Body fat (%) |  |
| Males | 18.6 ± 8.5 |
| Females | 28.2 ± 8.5 |
| ***Clinical characteristic*** |  |
| Systolic blood pressure (mmHg) | 115.2 ± 21.7 |
| Diastolic blood pressure (mmHg) | 77.3 ± 15.0 |

<sup>1</sup>Values are means ± SDs or medians [IQRs] unless otherwise indicated.

**Figure 7.**
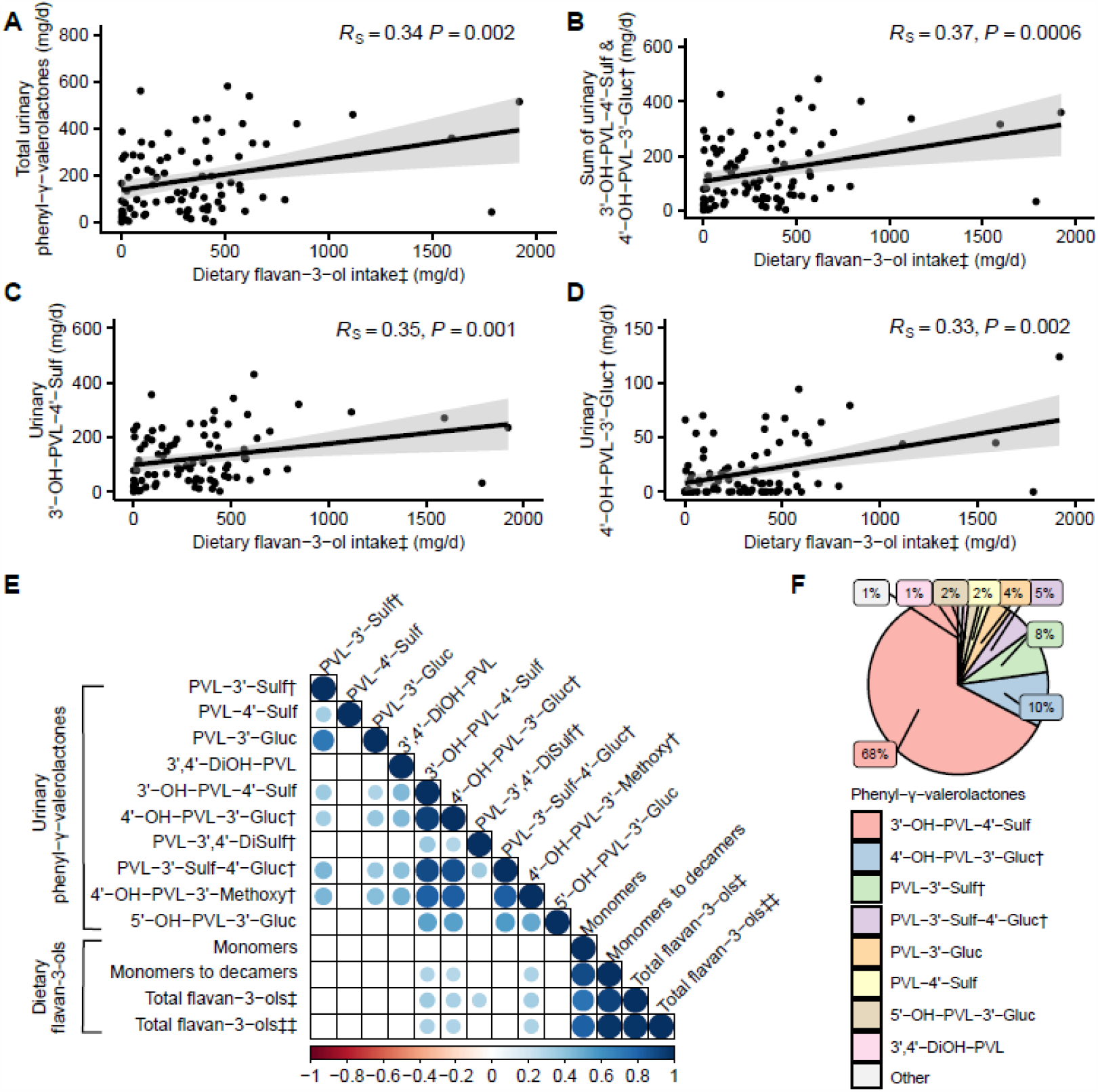
Relation between estimated dietary flavan-3-ol intake and 24-hour urinary phenyl-γ-valerolactone excretion in a cross-sectional study of free-living healthy participants (*n* = 86). Panels **A** through **D** show selected associations of interest, while panel **E** displays a correlation heatmap among flavan-3-ol sub-categories and all measured phenyl-γ-valerolactones. In the heatmap, blue circles show the strength of significant positive correlations and blank (white panels) are non-significant correlations. Panel **F** shows the average percent excretion of the measured phenyl-γ-valerolactones in the observational study. All correlations were calculated using Spearman’s correlation coefficient (*R*_*s*_). *P* < 0.01 was considered significant. Descriptive statistics of this analysis can be found in Supplementary Table 7. Structural information about the phenyl-γ-valerolactones is detailed in Supplementary Table 1. ^†^Tentatively identified compound. ‡Flavan-3-ol intake is the sum of (epi)catehins, their galloyl substituted derivatives plus proanthocyanidins. ‡‡Further includes (epi)gallocatehins and their galloyl substituted derivatives.

Examining subcategories of flavan-3-ol intake, the strength of the association became weaker when 3’
s-OH-PVL-4’-Sulf and 4’-OH-PVL-3’-Gluc were examined against the sum of monomers to decamers (3’-OH-PVL-4’-Sulf, *R*_s_ = 0.31, *P* = 0.004; 4’-OH-PVL-3’-Gluc, *R*_s_ = 0.28, *P* = 0.008), and the sum of monomers (3’-OH-PVL-4’-Sulf, *R*_s_ = 0.24, *P* = 0.02; 4’-OH-PVL-3’-Gluc, *R*_s_ = 0.22, *P* = 0.04). Correlations of other PVL compounds with dietary intake were also observed although, collectively, these compounds comprised <20% of total PVL excretion (Figure 7). When a heatmap of associations between PVLs was drawn, it was apparent that 3’-OH-PVL-4’-Sulf and tentatively identified 4’-OH-PVL-3’-Gluc, tended to associate the most frequently with other measured PVLs (Figure 7).

## Discussion

In a series of analyses, we assessed the performance of a range of PVLs as possible biomarkers of dietary flavan-3-ol exposure. Converging results from both studies revealed that two urinary PVLs [3’
s-OH-PVL-4’-Sulf and tentatively identified 4’-OH-PVL-3’-Gluc] appear as suitable biomarkers indicative of flavan-3-ol intake. These results were consistent, whether the compounds were measured in 24-hour urines or first morning voids. Overall, the identified PVLs, when used as biomarkers, either alone, or preferably in combination, may allow for the ranking of dietary flavan-3-ol exposure, in groups of individuals. Our findings will be useful to investigators conducting epidemiologic studies that examine potential relationships between food rich in flavan-3-ols and human health and disease.

As part of the current investigation, we measured a panel of 10 different PVLs to determine which may be the most suitable biomarkers of flavan-3-ol rich food intake. We found two specific PVLs [3’
s-OH-PVL-4’-Sulf and tentatively identified 4’-OH-PVL-3’-Gluc], accounted for most PVL excretion (≥75%) in our studies. This is in agreement with prior reports that show a select few PVLs, and in particular those 3’,4’-DiOH-PVLs with a sulfate or glucuronidate conjugate, are the principle metabolites derived, which constitute most of the circulating PVL pool, highlighting their candidacy as biomarkers for development (10–12). Direct evidence has shown 3’ and 4’ hydroxylate PVLs are derived from the breakdown of (epi)catechins and their gallate esters (as opposed to (epi)gallocatechins and their gallate esters (3)) which are known to occur in all of our RCT interventions (i.e., apple, cocoa, black tea and green tea) and to a lesser extent procyanidins (which are found in larger quantities in cocoa and apples but not in green or black tea) (2,13). Indeed, in the observational study, the sum of these compounds correlated with flavan-3-ol intake in a dose-dependent manner (*R*_s_ = 0.37) and in the RCT, their sum was significantly higher than the water (control) following each intervention. Interestingly, in the observational study both 3’-OH-PVL-4’-Sulf and 4’-OH-PVL-3’-Gluc showed associations with the sum of monomer to decamer intake (*R*_s_ = 0.28–0.31), the strength of which was not markedly improved when polymers were added to the calculation of flavan-3-ol exposure (*R*_s_ = 0.32–0.35). This suggests polymers do not form PVLs as readily as monomers, which supports prior findings (13). Moreover, in the RCT, when 3’-OH-PVL-4’-Sulf and 4’-OH-PVL-3’-Gluc were examined individually, there was a shift from sulfation towards glucuronidation, wherein the glucuronide became the more abundant form, with higher flavan-3-ol containing interventions. It is established that sulfation is a higher-affinity, lower-capacity pathway than glucuronidation, so when the dose of flavan-3-ol intake increases, a shift from sulfation toward glucuronidation occurs (14,15) which appears to account for these findings. This mechanism might also explain why 3’-OH-PVL-4’-Sulf accounted for a greater proportion of total PVL excretion in the observational study (68%) as compared to the RCT (44%), given the average flavan-3-ol intake in the observational study was less. The overall implications are, that measurement of both glucuronidated and sulfated derivatives may more accurately capture participants at the lower and higher ends of flavan-3-ol exposure. From these observations, we draw the conclusion that 3’-OH-PVL-4’-Sulf and tentatively identified 4’-OH-PVL-3’-Gluc, may act as biomarkers or a biosignature of flavan-3-ol exposure regardless of the source of (epi)catechins and their polymers. This is in line with Favari *et al*., who also highlighted the importance of these two PVLs as biomarkers of intake for cranberry flavan-3-ols (16).

Most PVLs measured throughout our studies were found in minor to moderate quantities; some of these PVLs merit comment. Firstly, in-line with prior reports, parent unconjugated PVLs were only present in modest quantities, with phase II metabolites predominating (2). Secondly, PVL-3’
s,4’-DiSulf and PVL-4’-Sulf were not detected in significantly higher quantities following any intervention, in comparison to the water control; Brindani *et al*., observed similar results following green tea intake (17). Thirdly, 5’-OH-PVL-3’-Gluc was only detected following consumption of green tea. Hydroxylated on the 3’ and 5’ positions of the B-ring, this PVL derivative forms exclusively from the breakdown of (epi)gallocatechins which are known to occur in high concentrations in green tea and in smaller concentrations in other foods such as nuts (e.g., almond, pecan etc.,), apples, berries (e.g., blackberries, blueberries etc.,) and avocados (2). As such, measurement of 5’-OH-PVL-3’-Gluc may be of benefit when investigating green tea consuming populations, although, measurement of a sulfated derivative may also be of value (18). To this end, population studies could benefit from a measuring a panel of PVLs which differentiate (epi)catechin vs. (epi)gallocatechin rich diets. Fourthly, of the observed mono-hydroxylated PVLs, those hydroxylated in the 3’ (as compared to 4’) position were found in greater quantities. Monagas *et al*., suggested that (epi)catechin dehydroxylation occurs preferentially at the 4’ position which could account for these findings (19). Overall, PVLs excreted in minor/moderate quantities do not appear to be the most suitable biomarkers for flavan-3-ol exposure, although, if (epi)gallocatechins are of interest, then researchers should consider measuring PVLs specifically derived from their breakdown. Moreover, minor PVL metabolites might also be key when assessing inter-individual variability in the production of PVLs (20,21).

Another major finding of the present investigation is that measurement of PVLs in first morning voided urine might be sufficient to rank flavan-3-ol exposure in groups of individuals. This is relevant to observational studies where 24-hour urine collections are rarely performed. We found the pattern of PVL excretion in the first morning spot urines almost mirrored the pattern of PVL excretion in the 24-hour urine. The main difference was the confidence intervals around the spot PVLs were wider. Consequently, we can conclude that spot urines are less sensitive than 24-hour urine measures. Although, our intervention timings were standardized to 9:00 am, 12:00 pm and 3:00 pm, and therefore, findings may have been different, if the treatments were consumed at random over the day’s duration. In the context of real-world food consumption, certain flavan-3-ol containing products, such as black tea, may be consumed more consistently over the day’s duration, whereas others, such as apples or drinking cocoa might be consumed on one occasion throughout the day—research to examine the impact of intake timing on first morning urine PVL status is therefore needed. Other researchers have also hypothesized that spot urines collected at random may also rank flavan-3-ol exposure (3), although evidence to confirm this hypothesis requires further study. Overall, while first morning spot urine samples might be adequate to rank flavan-3-ol exposure in large cohorts, 24-hour urine measures are superior and preferred when available.

We also evaluated PVL kinetics, following repeated days of flavan-3-ol intake, to examine how changes in habitual dietary intake may be reflected in the urine. We found that after two days of treatment, there was no accumulation of urinary PVL excretion, and following withdrawal of treatment on the third day, there was a return towards negligible PVL excretion. These findings are supported by known time-courses of PVL production, which tend to reach peak concentration around ∼4 to ∼20 hours post flavan-3-ol intake, before tapering to sustained lower production for up to 48 hours (10,18). The implications are, that urinary PVLs, when used as biomarkers, are most sensitive to a single day’s flavan-3-ol exposure. In the context of biomarker use in observational studies, if a participant is an irregular consumer, that happens to consume a high amount of flavan-3-ols on one specific day, they might be mistakenly classified as a regular consumer, whereas, if someone is a regular consumer, but does not consume a high intake for one certain day, they may be mistakenly classified as a low consumer. These findings compel us to conclude that repeated measures of PVL excretion, will allow for an improved indication of regular flavan-3-ol exposure. To this end, the use of questionnaires in conjunction with biomarkers is thought to provide a more thorough appraisal of true phenolic exposure (22) Further research is required to clearly elucidate the number of repeated measures needed to best approximate an individual’s habitual flavan-3-ol exposure, be it repeated 24-hour urine or spot samples.

Much of our work has indicated a beneficial impact of estimated dietary flavan-3-ol intake on CVD risk (23–27). This in alignment with the recent release of the first ever dietary bioactive guideline for flavan-3-ol intakes and cardiometabolic health (28). Most interestingly, the investigation of health outcomes utilizing PVLs as flavan-3-ol biomarkers has started to gain some traction. Ottaviani *et al*., recently measured PVL status in 25,618 participants from the European Prospective Investigation into Cancer (EPIC) Norfolk cohort (29). Spot urines were analyzed for 4’
s-OH-PVL-3’-Gluc and 4’-OH-PVL-3’-Sulf (i.e., isomers of the two predominate urinary PVL identified in the present work) and the sum was found to associate with lower blood pressure, but not long-term CVD risk (29). Urinary PVL status was also assessed in the recent COcoa Supplement and Multivitamin Outcomes Study (COSMOS) trial, although any association with CVD risk is yet to be reported (30). To our knowledge, only one other clinical trial has measured PVLs and examined changes in health status (31). Rodriguez-Mateos *et al*., fed participants cranberry juice and found that plasma 3’-OH-PVL-4’-Sulf (the predominate urinary PVL identified in the present work) significantly correlated with improved flow mediated dilation at 4 and 8 hours postprandial (31). Collectively, these findings are promising, although, work linking PVLs with health effects is still scarce; our results encourage the measurement of PVLs in future RCTs and observational studies.

This investigation had several limitations. While we examined a large range of PVL compounds as potential biomarkers for development, our panel was not exhaustive, and so, future work should evaluate other plausible candidates. Our study also relied on putative methods for the identification of several PVLs and for these structures, we can only speculate on their probable arrangement. For example, 4’
s-OH-PVL-3’-Gluc was isomerically identified using 4’-OH-PVL-5’-Gluc (for which we had a reference standard) and therefore we are less certain whether the glucuronide occurs on the 3’ or 4’ position of the B-ring, which has implications for biomarker development. Also, the absolute quantity of putative analytes cannot be definitively established, and these analytes may be under- or over-estimated in abundance (32). Quantitation, when possible, against authentic reference standards is recommended in future studies (32). Another notable observation was the scarcity of PVLs following black tea consumption. This likely reflects the structure of precursor flavan-3-ols found within our black tea intervention. Results obtained from black tea interventions containing higher levels of catechins should produce dose-dependent increments in PVLs. The broader implications are that the use of PVLs as biomarkers allows for the objective ranking of flavan-3-ol exposure, rather than an estimate from food composition databases, which may be inaccurate for certain products. Indeed, food-composition database limitations may also account for the modest correlations in the observational study, in addition to other factors such inter-individual variations in the host microbiota (33), which would only be observable in the RCT as consistently high or low PVL producing participants. It was also noticeable that the absolute quantity of urinary PVLs was quite high following cocoa treatment, although independent laboratory analysis of the cocoa product (using methods previously reported (34)), revealed a flavan-3-ol dose of ∼2000 mg/d (email communication), which indicates ∼50% conversion rate of flavan-3-ols to PVLs, which conforms with prior findings (11). Lastly, across our investigation, significance testing was also conducted at the 1% probability threshold, which some may consider too liberal. We consider our results best interpreted in the context of effects sizes, biological plausibility, sample power considerations, and hypothesis testing when considering whether certain PVLs merit consideration as biomarkers. Those compounds excreted in trace or minor amounts, may indeed be present in statistically discernable quantities, although they appear to represent such minor metabolic pathways, that their utility as biomarkers is not strongly supported.

In conclusion, urinary 3’
s-OH-PVL-4’-Sulf and tentatively identified 4’-OH-PVL-3’-Gluc are recommended as biomarkers for dietary flavan-3-ol exposure. These compounds reflect flavan-3-ol intake in a dose-dependent manner, and we demonstrated their capacity to rank flavan-3-ol consumption in a free-living population. Their measurement in first morning voids might be adequate to rank flavan-3-ol exposure in large cohorts, although 24-hour urine measures are preferred when available. Repeated measures of urinary PVL status are also recommended to best approximate an individual’s habitual flavan-3-ol exposure. The use of PVL as biomarkers indicative of flavan-3-ol exposure, and the potential protective benefits of flavan-3-ol rich diets, warrants further investigation.

## Data Availability

The data that support the findings of this study are available from the corresponding author upon reasonable request in-line with governing ethical considerations.

## Acknowledgements

NPB, KM & CK designed the research (project conception, development of overall research plan, and study oversight); BHP conducted the RCT; AG conducted the cross-sectional study; BHP calculated the flavonoid intake from food diaries; SS did the liquid chromatography tandem mass spectroscopy method development; BHP performed laboratory analysis; BHP analysed data; BHP wrote the paper; NPB and BHP had primary responsibility for final content; all authors critically reviewed the final draft of the manuscript. All authors read and approved the final version. We would like to thank Stella Kassara for valuable discussions concerning the contents of CocoaVia.

