## Supplemental File for "Performance of urinary phenyl-γ-valerolactones as biomarkers of dietary flavan-3-ol exposure"

Parmenter *et al.*

### Supplementary Methods

#### Clinical trial: Participant recruitment

Healthy men and women were recruited from a database of individuals who had participated in our previous studies and who had agreed to be contacted for future studies. Advertisements were also placed in local Newspapers. Before enrolment, participants were screened via telephone and then invited to the University of Western Australia, School of Biomedical Sciences, located at the Research Foundation Building of the Royal Perth Hospital for further assessment. After a standard medical and physical activity questionnaire, trained personal assessed anthropometry (height, weight and waist:hip circumference), conducted bio-impedance analysis (BMR, fat mass and fat free mass) and took blood pressure measurements (mean of 4 measures). Exclusion criteria were as follows: Age <18 or >80 years; body mass index <18.5 or >35 kg/m$^2$; systolic blood pressure ≥150 mmHg; diastolic BP ≥100 mmHg; current or recent (<12 months) smoking; diagnosed diabetes, cancer, psychiatric illness, cardiovascular disease, peripheral vascular disease or any other major illness; current use of any prescription medications or illicit drugs; use of antibiotics within previous 3 months; alcohol intake >210 g per week for women and >280 g per week for men; allergies to any study meal or intervention ingredients; gastro-intestinal tract disorders; previous gastro-intestinal surgery (except appendectomy); self-reported malabsorption (e.g. difficulty digesting or absorbing nutrients from food, potentially leading to bloating, cramping or gas); diarrhoea within the last 3 months; the use of a non-traditional diet (vegan, weight loss etc); use of dietary or herbal supplements; those currently participating in another clinical or dietary intervention study; and women who are lactating, pregnant or wishing to become pregnant during the study.

#### Clinical trial: Standardisation of dietary intake

Diets were prepared by the Nutrition Laboratory at Edith Cowan University. These were prepared using foods that contain no or very low flavan-3-ols contents. All participants consumed the same meals before each visit, apart from selected available snacks to raise energy intake according to individual appetite. Breakfast was a selection from three low-flavonoid meals: cornflakes with milk/rice krispies with milk/eggs on white toast. A low-flavonoid lunch was provided consisting of white bread or rice with either pumpkin soup or chicken and sweet corn soup. Dinner was a selection of: meat pie/mince samosas/chicken in pastry. Snacks consisted of the following options: custard cream biscuit/plain yoghurt/Babybel cheese/tinned pineapple/popcorn. The low (poly)phenolic diet contained less than three servings of fruit and vegetables per day. Water was provided ad labium.

**Analysis of urinary phenyl-γ-valerolactones**

Standards were kindly provided by Prof. Claudio Curti [University of Parma, Parma, Italy] (**Supplementary Table 1)** (1). The internal standard phloridzin (PZD) was purchased from Biochemika (Merck). All chemicals and solvents used for liquid chromatography tandem mass spectroscopy analysis (LC-MS/MS) were of LC-MS grade quality.

Urine (50 µL) was thawed on ice, centrifuged, and then transferred to a 96-well loading plate containing internal standards, PZD (5 µL) of [5 µg/mL] stock solution in methanol. After diluting with 100 µL 1% formic acid (FA) in water, the plate was covered with a seal mat and placed on ice and put on a microplate shaker for 2-5 mins. Samples were then loaded on to a Strata-X polymeric reversed phase micro elution 96-well plate, 2 mg/well (Phenomenex, Australia Pty Ltd.) that was pre-equilibrated with 200 µL (1% FA in methanol) and 200 µL (1% FA in water). The loading plate wells were rinsed with 100 µL (1% FA in water) and added to the Strata-X 96-well extraction plate with the samples and allowed to drain. Loaded samples were washed with 200 µL (1% FA in water) followed by 200 µL (0.1% FA in water). The PVLs were eluted with 100 µL (0.1% FA in methanol) into a 96-well collection plate containing 30 µL of 0.1% FA in water using a positive pressure manifold. The collection plates were sealed with aluminium foil and stored at -80°C for further LC-MS/MS analysis. All urine samples were done in duplicates.

The extracted PVLs were analysed on a Thermo Scientific TSQ Quantum Ultra Triple Quadrupole mass spectrometer equipped with a HESI source attached to an Accela High Performance liquid chromatography (HPLC) system. The mass spectrometer was operated in the negative ion multiple reaction monitoring (MRM) mode using argon as collision gas. Nitrogen was used as sheath, auxillary and sweep gas set to 50, 10 and 1 arbitrary units and the CID gas at 1.2 mTorr. The vaporizer temperature of ESI source was at 300 °C and spray voltage at 3 kV. The Q1 and Q3 mass resolution of the spectrometer was at 0.7 FWHM. The MRM transitions and MS optimization parameters monitored for the PVL standards, isomers and putatives are listed in **Supplementary Table 2**.

Compounds were separated by HPLC on a reverse-phase Kinetex EVO C18 column (2.6 μm, 100×2.1 mm) from Phenomenex. The mobile phases consisted of 0.1% formic acid in LCMS grade water (solvent A) and methanol containing 0.1% formic acid (solvent B) at a flow rate of 300 µL/min. The column and tray temperature of the Accela autosampler was maintained at 35°C and 10°C, respectively. The run time for the LC method was 16 mins and the gradient conditions were as follows, 5% B at 0-0.5 mins, increased to 95% B at 10 mins, held at 95% B to 13 mins, reduced to 5% B at 13.01 mins and held at 5% B to 16 mins to equilibrate to starting conditions.

Concentrations were determined using calibration curves for each of the phenyl-γ-valerolactones by spiking "*stripped*" urines with available standards at different dilutions. This was a pooled sample and polyphenols were extracted by passage through a column of polyvinylpolypyrrolidone. Linear regression analysis, of ratio of area of each compound to internal standard, PZD, as a function of varying concentration of PVLs typically gave $R^2$ >

0.90-0.99, were used for quantitative analysis in ng/mL. The LOD and LOQ was 50 ng and 100 ng respectively. The % CV for all measured PVLs was 5-15%. Data acquisition was performed with Xcalibur (2.0.7 version) and data analysis was done using QuanBrowser, Thermo Xcalibur (version 4.1.50).

For quantitative analysis of isomers and putatives, different volumes (25 µL- 100 µL) of a pooled urine sample (high polyphenolic diet) were extracted under similar conditions using the Strata-X micro elution 96-well plate method as described above. Linear regression analysis used ratio of area of isomers/putatives to internal standard (PZD) as a function of concentration of the isomer/putative estimated from the response factor of calibration curves for available standards. The standard for each isomer and putative was selected based on their chemical structure. For example, linear regression fit for isomer 4′-OH-PVL-3′-Gluc was carried out using the regression analysis fit for known standard 5′-OH-PVL-3′-Gluc. Similarly, concentrations for 4′-OH-PVL-3′-Methoxy, PVL-3′,4′-DiSulf, and PVL-3′-Sulf-4′-Gluc were calculated using the regression fits of known standards 3′,4′-DiOH-PVL, PVL-4′-Sulf and 5′-OH-PVL-3′-Gluc respectively.

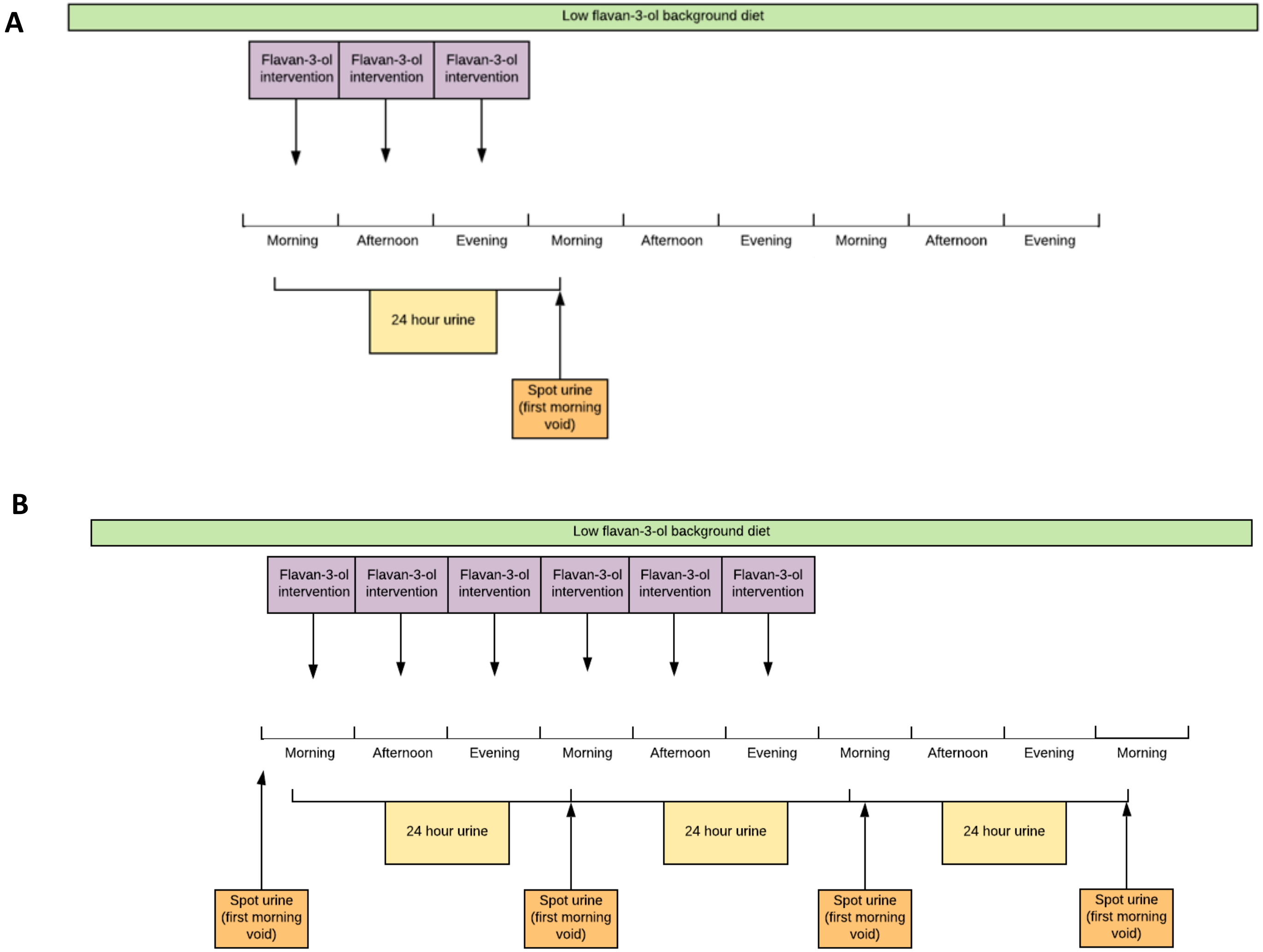


**Supplementary Figure 1.** Schematic of the 5-way randomised clinical trial. Panel **A** shows the testing procedure for the regular intervention periods. Panel **B** shows testing procedure for the extended intervention period.

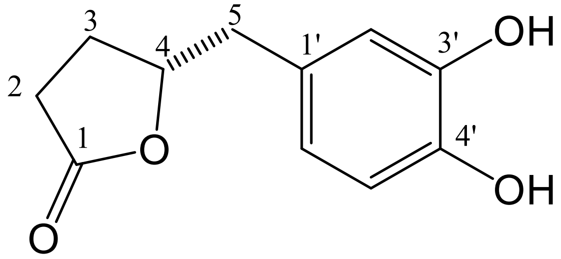


**Supplementary Figure 2.** Nomenclature and numbering of phenyl-γ-valerolactones metabolites.

**Supplementary table 1**. Nomenclature and structural identification of the phenyl-γ-valerolactones measured in the study

| Name | Abbreviations | Alternative possible structural arrangements†† |
|---|---|---|
| 5-Phenyl-γ-valerolactone-3′-sulfate* | PVL-3′-Sulf | 5-Phenyl-γ-valerolactone-5′-sulfate* |
| 5-Phenyl-γ-valerolactone-4′-sulfate‡ | PVL-4′-Sulf | n.a. |
| 5-Phenyl-γ-valerolactone-3′-glucuronide‡ | PVL-3′-Gluc | n.a. |
| 5-(3′,4′-Dihydroxyphenyl)-γ-valerolactone‡ | 3′,4′-DiOH-PVL | n.a. |
| 5-(3′-Hydroxyphenyl)-γ-valerolactone-4′-sulfate ‡ | 3′-OH-PVL-4′-Sulf | n.a. |
| 5-(4′-Hydroxyphenyl)-γ-valerolactone-3′-glucuronide* | 4′-OH-PVL-3′-Gluc | 5-(3′-Hydroxyphenyl)-γ-valerolactone-4′-glucuronide* |
| 5-Phenyl-γ-valerolactone-3′,4′-disulfate† | PVL-3′,4′-DiSulf | 5-Phenyl-γ-valerolactone-3′,5′-disulfate† |
| 5-Phenyl-γ-valerolactone-3′-sulfate,4′-glucuronide† | PVL-3′-Sulf-4′-Gluc | 5-Phenyl-γ-valerolactone-4′-sulfate,3′-glucuronide†<br>5-Phenyl-γ-valerolactone-3′-sulfate,5′-glucuronide† |
| 5-(4′-Hydroxyphenyl)-γ-valerolactone-3′-methoxy† | 4′-OH-PVL-3′-Methoxy | 5-(3′-Hydroxyphenyl)-γ-valerolactone-4′-methoxy † |
| 5-(5′-Hydroxyphenyl)-γ-valerolactone-3′-glucuronide‡ | 5′-OH-PVL-3′-Gluc | n.a. |

‡Compounds identified with reference standards

*Possible structures of compounds identified as isomers of those with reference standards

†Possible structures of compounds putatively identified (without reference standard)

††Our study relied on putative methods for the identification of several PVLs and for these structures, we can only speculate on their probable arrangement. This column shows alternative possible structural arrangements, for the putatively identified compounds. Putatively identified compounds, were measured as single compounds, and do not reflect the measurement of several structures.

**Supplementary table 2**. MRM transitions, MS optimization parameters and RT for phenyl-γ-valerolactones

| Compound | Parent *m/z* ion | Product *m/z* ions | Collision Energy (eV) | RT (min) |
|---|---|---|---|---|
| PVL-4′-Sulf | 271 | 191 (QF), 147 | 23(QF), 35 | 5.7 |
| 3′-OH-PVL-4′-Sulf | 287 | 207 (QF), 163 | 23 (QF), 34 | 5.4 |
| 5′-OH-PVL-3′-Gluc | 383 | 207 (QF), 163 | 23 (QF), 35 | 3.3 |
| PVL-3′-Gluc | 367 | 191 (QF), 147 | 23 (QF), 35 | 4.4 |
| 3′,4′-DiOH-PVL | 207 | 163 (QF), 122 | 20 (QF), 24 | 4.3 |
| 4′-OH-PVL-3′-Methoxy | 221 | 206 (QF) | 20 (QF) | 6.3 |
| PVL-3′,4′-DiSulf | 367.1 | 287 (QF), 243 | 21(QF), 24 | 6.6 |
| PVL-3′-Sulf-4′-Gluc | 463 | 287 (QF), 383 | 23 (QF), 20 | 4.8 |

RT, retention time; QF, quantifier ion.

### Supplementary Results

**Supplementary Table 3.** Excretion of phenyl-γ-valerolactones in 24-hour urines following flavan-3-ol containing interventions of either cocoa, apple, green tea, black tea or a water control in a 5-way randomised cross over trial (n = 16).[1]

| | Water | Black tea | | Green tea | | Apple | | Cocoa | |
|---|---|---|---|---|---|---|---|---|---|
| Metabolite (mg/d) | Median [IQR] | Median [IQR] | *P*‡ | Median [IQR] | *P*‡ | Median [IQR] | *P*‡ | Median [IQR] | *P*‡ |
| PVL-3′-Sulf† | 0.0 [0.0, 0.0] | 0.0 [0.0, 0.3] | 0.5322 | 33.2 [8.0, 65.5] | <0.0001 | 8.5 [0.6, 28.4] | <0.0001 | 55.7 [19.7, 96.0] | <0.0001 |
| PVL-4′-Sulf | 0.0 [0.0, 0.0] | 0.0 [0.0, 0.0] | 0.8821 | 0.0 [0.0, 0.0] | 0.2113 | 0.0 [0.0, 0.0] | 0.3974 | 0.0 [0.0, 0.0] | 0.228 |
| PVL-3′-Gluc | 0.0 [0.0, 0.0] | 0.0 [0.0, 0.0] | 0.9108 | 29.1 [4.4, 48.2] | <0.0001 | 2.1 [0.0, 12.2] | <0.0001 | 60.5 [24.1, 139.8] | <0.0001 |
| 3′,4′-DiOH-PVL | 0.3 [0.3, 0.4] | 0.2 [0.1, 0.3] | 0.7194 | 2.5 [1.4, 3.0] | <0.0001 | 1.1 [0.7, 1.5] | 0.0317 | 20.2 [12.4, 30.3] | <0.0001 |
| 3′-OH-PVL-4′-Sulf | 1.0 [0.6, 1.2] | 14.7 [9.8, 22.1] | <0.0001 | 141.7 [119.6, 173.2] | <0.0001 | 116.6 [93.6, 147.7] | <0.0001 | 441.8 [343.5, 555.2] | <0.0001 |
| 4′-OH-PVL-3′-Gluc† | 0.0 [0.0, 0.0] | 0.0 [0.0, 0.0] | 0.9975 | 65.4 [43.8, 91.2] | <0.0001 | 18.1 [0.5, 24.6] | <0.0001 | 483.4 [362.2, 655.6] | <0.0001 |
| PVL-3′,4′-DiSulf† | 0.0 [0.0, 0.0] | 0.1 [0.0, 0.4] | 0.4167 | 0.0 [0.0, 0.1] | 0.7621 | 0.0 [0.0, 0.0] | 0.9255 | 0.2 [0.0, 0.5] | 0.3044 |
| PVL-3′-Sulf-4′-Gluc† | 0.0 [0.0, 0.0] | 0.0 [0.0, 0.0] | 0.8906 | 4.3 [1.9, 11.9] | <0.0001 | 2.5 [1.3, 12.3] | <0.0001 | 25.5 [17.8, 39.7] | <0.0001 |
| 4′-OH-PVL-3′-Methoxy† | 0.0 [0.0, 0.0] | 0.0 [0.0, 0.1] | 0.887 | 0.5 [0.4, 0.7] | 0.0381 | 0.4 [0.3, 0.5] | 0.1241 | 3.3 [2.5, 4.5] | <0.0001 |
| 5′-OH-PVL-3′-Gluc | 0.3 [0.3, 0.5] | 0.7 [0.5, 1.0] | 0.2689 | 29.7 [21.1, 48.6] | <0.0001 | 0.3 [0.1, 0.4] | 0.9564 | 0.3 [0.2, 0.5] | 0.833 |
| Sum of 3′-OH-PVL-4′-Sulf & 4′-OH-PVL-3′-Gluc† | 1.0 [0.6, 1.2] | 14.7 [9.8, 22.1] | <0.0001 | 212.2 [168.0, 259.9] | <0.0001 | 149.1 [103.8, 169.0] | <0.0001 | 909.1 [805.6, 1070.4] | <0.0001 |
| Total phenyl-γ-valerolactones | 1.6 [1.4, 2.6] | 17.0 [11.5, 23.5] | <0.0001 | 289.4 [269.3, 456.6] | <0.0001 | 166.3 [115.4, 216.1] | <0.0001 | 1151.8 [974.6, 1266.4] | <0.0001 |

[1]Differences in total phenyl-γ-valerolactone excretion across interventions was assessed by linear mixed model on log-transformed data, when applicable.

‡*P*-value for the comparison of treatment to water. $P < 0.01$ was considered significant.

†Tentatively identified compound.

**Supplementary Table 4.** Excretion of phenyl-γ-valerolactones in 24-hour urines following repeated days of flavan-3-ol containing interventions (on days one and two), preceding withdrawal of treatment (on the third day), collected during the extended RCT intervention period (cocoa *n* = 2; apple *n* = 4; green tea *n* = 3; black tea *n* = 3; water *n* = 3).[1]

| | Phenyl-γ-valerolactones (mg/d) | | | |
|---|---|---|---|---|
| | 3′-OH-PVL-4′-Sulf | 4′-OH-PVL-3′-Gluc† | Sum of 3′-OH-PVL-4′-Sulf & 4′-OH-PVL-3′-Gluc† | Total phenyl-γ-valerolactones |
| Water | | | | |
| Day 1 | 0.6 [0.6, 0.6] | 0.0 [0.0, 0.0] | 0.6 [0.6, 0.6] | 1.2 [1.1, 1.3] |
| Day 2 | 0.6 [0.5, 0.7] | 0.0 [0.0, 0.0] | 0.6 [0.5, 0.7] | 1.1 [1.0, 1.4] |
| *P*‡ | 0.9490 | 0.9999 | 0.9397 | 0.9828 |
| Day 3 | 0.5 [0.5, 0.7] | 0.0 [0.0, 0.0] | 0.5 [0.5, 0.7] | 1.1 [1.1, 1.2] |
| *P*‡‡ | 0.9995 | 0.9999 | 0.9994 | 0.9461 |
| *P*‡‡‡ | 0.9485 | 0.9999 | 0.9391 | 0.9290 |
| Black tea | | | | |
| Day 1 | 9.8 [7.7, 22.4] | 0.0 [0.0, 0.0] | 9.8 [7.7, 22.4] | 10.7 [8.4, 24.8] |
| Day 2 | 10.5 [8.2, 25.9] | 0.0 [0.0, 0.1] | 10.5 [8.2, 26.0] | 12.9 [9.8, 29.9] |
| *P*‡ | 0.8346 | 0.8948 | 0.8021 | 0.7171 |
| Day 3 | 2.2 [1.6, 3.5] | 0.0 [0.0, 0.0] | 2.2 [1.6, 3.5] | 2.9 [2.5, 4.8] |
| *P*‡‡ | 0.0009 | 0.9999 | 0.0006 | 0.0047 |
| *P*‡‡‡ | 0.0004 | 0.8948 | 0.0003 | 0.0020 |
| Green tea | | | | |
| Day 1 | 138.5 [122.1, 148.5] | 51.8 [46.6, 57.7] | 189.6 [168.0, 206.2] | 281.7 [255.4, 286.0] |
| Day 2 | 103.9 [93.2, 116.1] | 31.5 [23.1, 38.2] | 135.3 [122.7, 148.0] | 186.9 [147.0, 220.5] |
| *P*‡ | 0.5236 | 0.0691 | 0.2857 | 0.2564 |
| Day 3 | 3.7 [1.7, 6.4] | 0.0 [0.0, 0.0] | 3.7 [1.7, 6.4] | 4.3 [2.0, 7.6] |
| *P*‡‡ | <0.0001 | <0.0001 | <0.0001 | <0.0001 |
| *P*‡‡‡ | <0.0001 | <0.0001 | <0.0001 | <0.0001 |
| Apple | | | | |
| Day 1 | 149.5 [122.1, 191.1] | 12.5 [0.3, 30.1] | 172.7 [127.8, 226.4] | 185.8 [138.2, 257.3] |
| Day 2 | 114.2 [81.9, 133.2] | 1.8 [0.0, 11.5] | 126.8 [84.7, 152.7] | 150.6 [104.5, 172.3] |
| *P*‡ | 0.1864 | 0.1107 | 0.1130 | 0.1554 |
| Day 3 | 4.0 [2.1, 13.9] | 0.0 [0.0, 0.0] | 4.0 [2.1, 13.9] | 5.1 [3.1, 19.8] |
| *P*‡‡ | <0.0001 | <0.0001 | <0.0001 | <0.0001 |
| *P*‡‡‡ | <0.0001 | 0.0004 | <0.0001 | <0.0001 |
| Cocoa | | | | |
| Day 1 | 342.3 [300.2, 384.4] | 319.2 [284.0, 354.4] | 661.5 [584.3, 738.8] | 991.5 [864.0, 1119.1] |
| Day 2 | 300.6 [270.7, 330.5] | 388.6 [369.7, 407.6] | 689.2 [640.4, 738.1] | 1057.6 [937.6, 1177.5] |
| *P*‡ | 0.8165 | 0.6720 | 0.8916 | 0.8787 |
| Day 3 | 84.7 [77.4, 92.1] | 23.1 [21.9, 24.4] | 107.9 [99.3, 116.4] | 164.9 [144.9, 185.0] |
| *P*‡‡ | 0.0076 | <0.0001 | 0.0004 | 0.0009 |
| *P*‡‡‡ | 0.0146 | <0.0001 | 0.0003 | 0.0006 |

[1]Data presented as Median [IQR] unless stated. Differences in total phenyl-γ-valerolactone excretion across days was assessed by linear mixed model on log-transformed data, when applicable. Data not shown for phenyl-γ-valerolactones excreted in moderate to minor quantities.

‡*P*-value for the comparison of day one to day two.

‡‡*P*-value for the comparison of day two to day three.

‡‡‡*P*-value for the comparison of day one to day three.

†Tentatively identified compound.

**Supplementary Table 5.** Excretion of phenyl-γ-valerolactones in first morning spot urine samples following flavan-3-ol containing interventions of either cocoa, apple, green tea, black tea or a water control in a 5-way randomised cross over trial (n = 16).[1]

| Metabolite | Water | Black tea | | Green tea | | Apple | | Cocoa | |
|---|---|---|---|---|---|---|---|---|---|
| (mg analyte per g creatinine) | Median [IQR] | Median [IQR] | *P*‡ | Median [IQR] | *P*‡ | Median [IQR] | *P*‡ | Median [IQR] | *P*‡ |
| PVL-3′-Sulf† | 0.0 [0.0, 0.0] | 0.2 [0.0, 0.7] | 0.1161 | 53.3 [15.6, 116.3] | <0.0001 | 3.6 [1.3, 25.3] | <0.0001 | 119.6 [40.2, 206.1] | <0.0001 |
| PVL-4′-Sulf | 0.0 [0.0, 0.0] | 0.1 [0.1, 0.3] | 0.5513 | 0.0 [0.0, 0.0] | 0.9473 | 0.0 [0.0, 0.1] | 0.9543 | 0.0 [0.0, 0.0] | 0.973 |
| PVL-3′-Gluc | 0.0 [0.0, 0.0] | 0.0 [0.0, 0.0] | 0.6857 | 30.3 [8.5, 58.4] | <0.0001 | 0.6 [0.0, 6.4] | <0.0001 | 89.3 [40.6, 152.8] | <0.0001 |
| 3′,4′-DiOH-PVL | 0.0 [0.0, 0.0] | 0.0 [0.0, 0.0] | 0.9329 | 0.0 [0.0, 5.5] | 0.0003 | 0.0 [0.0, 0.0] | 0.999 | 34.0 [16.9, 47.0] | <0.0001 |
| 3′-OH-PVL-4′-Sulf | 1.6 [1.1, 2.8] | 31.7 [17.8, 57.2] | <0.0001 | 273.1 [160.0, 357.8] | <0.0001 | 138.7 [85.1, 194.3] | <0.0001 | 741.0 [491.8, 1121.0] | <0.0001 |
| 4′-OH-PVL-3′-Gluc† | 0.0 [0.0, 0.0] | 0.0 [0.0, 0.4] | 0.2435 | 108.9 [43.1, 175.1] | <0.0001 | 7.9 [0.3, 18.7] | <0.0001 | 666.9 [476.1, 1036.2] | <0.0001 |
| PVL-3′,4′-DiSulf† | 0.0 [0.0, 0.0] | 0.0 [0.0, 0.0] | 0.9349 | 0.0 [0.0, 0.0] | 0.8755 | 0.0 [0.0, 0.0] | 0.999 | 0.0 [0.0, 0.0] | 0.9177 |
| PVL-3′-Sulf-4′-Gluc† | 0.0 [0.0, 0.0] | 0.0 [0.0, 0.4] | 0.0393 | 22.0 [8.0, 28.0] | <0.0001 | 8.1 [2.6, 12.2] | <0.0001 | 88.9 [49.2, 118.9] | <0.0001 |
| 4′-OH-PVL-3′-Methoxy† | 0.0 [0.0, 0.0] | 0.0 [0.0, 0.0] | 0.8897 | 0.9 [0.4, 1.1] | 0.0162 | 0.2 [0.1, 0.4] | 0.3337 | 3.8 [2.6, 6.0] | <0.0001 |
| 5′-OH-PVL-3′-Gluc | 1.0 [0.7, 1.2] | 1.7 [1.5, 3.1] | 0.0277 | 65.7 [34.1, 177.0] | <0.0001 | 1.0 [0.7, 1.1] | 0.9054 | 0.7 [0.0, 1.2] | 0.418 |
| Sum of 3′-OH-PVL-4′-Sulf & 4′-OH-PVL-3′-Gluc† | 1.6 [1.1, 2.8] | 32.3 [18.0, 59.2] | <0.0001 | 438.1 [203.1, 538.2] | <0.0001 | 171.2 [89.1, 210.0] | <0.0001 | 1414.7 [971.5, 2107.6] | <0.0001 |
| Total phenyl-γ-valerolactones | 3.4 [2.2, 3.9] | 36.4 [20.3, 64.5] | <0.0001 | 636.1 [419.2, 1030.4] | <0.0001 | 192.2 [96.4, 240.7] | <0.0001 | 1900.5 [1360.3, 2552.8] | <0.0001 |

[1]Differences in total phenyl-γ-valerolactone excretion across interventions was assessed by linear mixed model on log-transformed data, when applicable.

‡*P*-value for the comparison of treatment to water. $P < 0.01$ was considered significant.

†Tentatively identified compound.

**Supplementary Table 6.** Excretion of phenyl-γ-valerolactones in first morning spot urines following repeated days of flavan-3-ol containing interventions (on days one and two), preceding withdrawal of treatment (on the third day), collected during the during the extended RCT intervention period (cocoa *n* = 2; apple *n* = 4; green tea *n* = 3; black tea *n* = 3; water *n* = 3).[1]

| | Phenyl-γ-valerolactones (mg analyte per g creatinine) | | | |
|---|---|---|---|---|
| | 3′-OH-PVL-4′-Sulf | 4′-OH-PVL-3′-Gluc† | Sum of 3′-OH-PVL-4′-Sulf & 4′-OH-PVL-3′-Gluc† | Total phenyl-γ-valerolactones |
| Water | | | | |
| Day 1 | 2.0 [1.8, 2.6] | 0.0 [0.0, 0.0] | 2.0 [1.8, 2.6] | 3.3 [3.1, 3.8] |
| Day 2 | 1.0 [1.0, 1.5] | 0.0 [0.0, 0.0] | 1.0 [1.0, 1.5] | 1.8 [1.7, 2.6] |
| Day 3 | 2.6 [2.3, 2.9] | 0.0 [0.0, 0.0] | 2.6 [2.3, 2.9] | 3.9 [3.3, 4.0] |
| Day 4 | 1.7 [1.6, 1.8] | 0.0 [0.0, 0.0] | 1.7 [1.6, 1.8] | 3.2 [2.9, 3.3] |
| *P* (D1 vs. D2) | 0.4656 | 0.9999 | 0.4983 | 0.4803 |
| *P* (D1 vs. D3) | 0.8416 | 0.9999 | 0.8524 | 0.9619 |
| *P* (D1 vs. D4) | 0.7169 | 0.9999 | 0.7359 | 0.8326 |
| *P* (D2 vs. D3) | 0.3528 | 0.9999 | 0.3895 | 0.4515 |
| *P* (D2 vs. D4) | 0.7134 | 0.9999 | 0.7326 | 0.6197 |
| *P* (D3 vs. D4) | 0.5739 | 0.9999 | 0.6013 | 0.7956 |
| Black tea | | | | |
| Day 1 | 2.0 [1.5, 5.5] | 0.0 [0.0, 0.0] | 2.0 [1.5, 5.5] | 4.2 [3.0, 7.6] |
| Day 2 | 25.6 [14.3, 41.1] | 0.0 [0.0, 0.0] | 25.6 [14.3, 41.1] | 28.5 [16.0, 43.5] |
| Day 3 | 29.5 [17.5, 31.1] | 0.0 [0.0, 0.5] | 29.5 [17.5, 31.6] | 31.7 [19.0, 34.7] |
| Day 4 | 1.5 [1.4, 2.0] | 0.0 [0.0, 0.0] | 1.5 [1.4, 2.0] | 3.0 [2.3, 3.0] |
| *P* (D1 vs. D2) | 0.0014 | 0.9999 | 0.005 | 0.0099 |
| *P* (D1 vs. D3) | 0.0011 | 0.6479 | 0.0041 | 0.0077 |
| *P* (D1 vs. D4) | 0.4369 | 0.9999 | 0.4708 | 0.3337 |
| *P* (D2 vs. D3) | 0.9493 | 0.6479 | 0.9388 | 0.9202 |
| *P* (D2 vs. D4) | 0.0001 | 0.9999 | 0.0007 | 0.0008 |
| *P* (D3 vs. D4) | 0.0001 | 0.6479 | 0.0006 | 0.0006 |
| Green tea | | | | |
| Day 1 | 2.9 [2.0, 3.6] | 0.0 [0.0, 0.0] | 2.9 [2.0, 3.6] | 4.2 [3.3, 5.3] |
| Day 2 | 368.1 [270.9, 416.9] | 206.8 [117.7, 274.9] | 611.3 [388.6, 728.2] | 862.0 [547.2, 1086.3] |
| Day 3 | 291.6 [214.8, 346.2] | 125.8 [53.2, 188.1] | 432.1 [290.9, 526.1] | 620.7 [405.7, 743.9] |
| Day 4 | 3.5 [2.6, 4.6] | 0.0 [0.0, 0.0] | 3.5 [2.6, 4.6] | 4.6 [3.6, 6.1] |
| *P* (D1 vs. D2) | <0.0001 | <0.0001 | <0.0001 | <0.0001 |
| *P* (D1 vs. D3) | <0.0001 | <0.0001 | <0.0001 | <0.0001 |
| *P* (D1 vs. D4) | 0.69 | 0.9999 | 0.7107 | 0.8361 |
| *P* (D2 vs. D3) | 0.6184 | 0.3869 | 0.532 | 0.3007 |
| *P* (D2 vs. D4) | <0.0001 | <0.0001 | <0.0001 | <0.0001 |
| *P* (D3 vs. D4) | <0.0001 | <0.0001 | <0.0001 | <0.0001 |
| Apple | | | | |
| Day 1 | 4.3 [1.6, 21.4] | 0.0 [0.0, 0.0] | 4.3 [1.6, 21.4] | 5.6 [2.7, 31.7] |
| Day 2 | 94.1 [45.3, 191.2] | 0.3 [0.0, 8.6] | 110.3 [45.3, 215.7] | 122.4 [48.9, 244.8] |
| Day 3 | 81.2 [68.2, 188.4] | 1.1 [0.2, 4.2] | 87.7 [71.2, 195.1] | 105.7 [87.2, 222.3] |
| Day 4 | 4.5 [2.4, 10.5] | 0.0 [0.0, 0.0] | 4.5 [2.4, 10.5] | 6.3 [3.5, 17.6] |
| *P* (D1 vs. D2) | <0.0001 | 0.0164 | <0.0001 | <0.0001 |
| *P* (D1 vs. D3) | <0.0001 | 0.0242 | <0.0001 | <0.0001 |
| *P* (D1 vs. D4) | 0.698 | 0.9999 | 0.7181 | 0.8089 |
| *P* (D2 vs. D3) | 0.8491 | 0.8826 | 0.886 | 0.8466 |
| *P* (D2 vs. D4) | <0.0001 | 0.0164 | <0.0001 | <0.0001 |
| *P* (D3 vs. D4) | <0.0001 | 0.0242 | <0.0001 | <0.0001 |
| Cocoa | | | | |
| Day 1 | 18.5 [10.5, 26.5] | 0.0 [0.0, 0.0] | 18.5 [10.5, 26.5] | 22.3 [14.0, 30.5] |
| Day 2 | 630.2 [630.2, 630.2] | 608.8 [608.8, 608.8] | 619.5 [309.7, 929.2] | 1900.5 [1900.5, 1900.5] |
| Day 3 | 417.8 [332.5, 503.2] | 486.9 [467.5, 506.3] | 904.7 [800.0, 1009.5] | 1287.7 [1192.7, 1382.6] |
| Day 4 | 15.5 [11.2, 19.8] | 0.0 [0.0, 0.0] | 15.5 [11.2, 19.8] | 23.3 [18.1, 28.4] |
| *P* (D1 vs. D2) | <0.0001 | <0.0001 | <0.0001 | <0.0001 |
| *P* (D1 vs. D3) | <0.0001 | <0.0001 | <0.0001 | <0.0001 |
| *P* (D1 vs. D4) | 0.6963 | 0.999 | 0.7165 | 0.6198 |
| *P* (D2 vs. D3) | 0.5145 | 0.784 | 0.8804 | 0.7469 |
| *P* (D2 vs. D4) | <0.0001 | <0.0001 | <0.0001 | <0.0001 |
| *P* (D3 vs. D4) | <0.0001 | <0.0001 | <0.0001 | <0.0001 |

[1]Data presented as Median [IQR] unless stated. Differences in total phenyl-γ-valerolactone excretion across days was assessed by linear mixed model on log-transformed data, when applicable. Data not shown for phenyl-γ-valerolactones excreted in moderate to minor quantities.

†Tentatively identified compound.

**Supplementary Table 7.** Estimated dietary flavan-3-ol intake and 24-hour urinary phenyl-γ-valerolactone excretion in a cross-section of free-living healthy participants (*n* = 86).[1]

| | All participants (n = 86) |
|---|---|
| Dietary intake | |
| Energy (kJ) | |
| Total flavan-3-ols‡ (mg/d) | 250.7 [72.7–409.7] |
| Total flavan-3-ols‡‡(mg/d) | 290.0 [78.3–460.3] |
| Monomers | 26.3 [8.0–100.8] |
| Monomers to decamers | 176.8 [43.7–328.8] |
| Polymers | 66.2 [15.3–136.5] |
| Urinary excretion | |
| Total phenyl-γ-valerolactones (mg/d) | 135 [53.5–283.2] |
| Mono-hydroxy-phenyl-γ-valerolactones | |
| 5-Phenyl-γ-valerolactone-3′-sulfate† | 4.8 [1.9–13.7] |
| 5-Phenyl-γ-valerolactone-4′-sulfate | 2.4 [0.3–5.0] |
| 5-Phenyl-γ-valerolactone-3′-glucuronide | 0 [0–4.3] |
| Di-hydroxy-phenyl-γ-valerolactones | |
| 5-(3′,4′-Dihydroxyphenyl)-γ-valerolactone | 0 [0–1.8] |
| 5-(3′-Hydroxyphenyl)-γ-valerolactone-4′-sulfate | 91 [41.2–198.0] |
| 5-(4′-Hydroxyphenyl)-γ-valerolactone-3′-glucuronide† | 6.0 [0–26.5] |
| 5-Phenyl-γ-valerolactone-3′,4′-disulfate† | 0.1 [0–1.2] |
| 5-Phenyl-γ-valerolactone-3′-sulfate,4′-glucuronide† | 5.3 [2.3–12.9] |
| 5-(4′-Hydroxyphenyl)-γ-valerolactone-3′-methoxy† | 0.3 [0.1–0.5] |
| 5-(5′-Hydroxyphenyl)-γ-valerolactone-3′-glucuronide | 1.1 [0.5–2.3] |

[1]Values are means ± SDs or medians [IQRs] unless otherwise indicated.

†Tentatively identified compound.

‡Flavan-3-ol intake is the sum of (epi)catehins, their galloyl substituted derivatives plus proanthocyanidins.

‡‡Additionally includes (epi)gallocatehins and their galloyl substituted derivatives.
